# Feasibility and cross-cultural validation of an adapted social skills group training programme (KONTAKT™ CHILD) for Chinese autistic children: a wait-list RCT protocol

**DOI:** 10.1101/2023.10.30.23296686

**Authors:** Uchong Lao, Huilin Zhu, Fengjing Liang, Enid Wuxia Bai, Peipei Yin, Xiaoqian Huang, Sonya Girdler, Sven Bölte, Xiaobing Zou

**Affiliations:** Child Development and Behavior Center, The Third Affiliated Hospital, Sun Yat-Sen University, China, 2693 Kai Chuang Avenue, Huangpu District, Guangzhou, 510530, China; Curtin Autism Research Group, School of Allied Health, Curtin University, Kent Street, Bentley, Perth, Western Australia; Center of Neurodevelopmental Disorders (KIND), Centre for Psychiatry Research, Division of Neuropsychiatry, Department of Women’s and Children’s Health, Karolinska Institutet & Stockholm Health Care Services, Region Stockholm; Child and Adolescent Psychiatry, Stockholm Health Care Services, Region Stockholm, Stockholm, Sweden

**Keywords:** Social skills group training, Children, KONTAKT™, Cross-cultural adaptation, Feasibility, Autism, Chinese

## Abstract

**Introduction:** School-aged autistic children commonly experience social communication and interaction challenges in their everyday lives. While international evidence suggests that social skills group training (SSGT) programmes can support autistic children, improving their psycho-social functioning, to date there is no standardised evidence-based SSGT tailored towards the needs of autistic children aged 8 to 12 years living in the Chinese Mainland. Therefore, the primary objective is to evaluate the feasibility and acceptability of a culturally adapted 16-session version of the social skills programme KONTAKT™ in Chinese autistic children. Additionally, preliminary efficacy and cost-effectiveness will be assessed to inform a future definitive trial.

**Methods and analysis:** This study will employ a randomised, single-blinded, wait-list controlled feasibility design with 36 Chinese autistic children aged 8 to 12 years with Intelligence Quotient over 70 referred to the Child Development Behaviour Centre, in Guangzhou, China. Participants will be randomly assigned to either the immediate training group or the delayed training group stratified by sex. Feasibility will be assessed using quantitative and qualitative data obtained from the KONTAKT™ CHILD participants, their parents, and facilitators of the programme. Preliminary efficacy and cost-effectiveness are assessed via quantitative data obtained at five time points. The primary efficacy outcome is participants’ improvement in social skills as measured by the Contextual

Assessment of Social Skills (CASS). Other outcomes include parents’ and teachers’ reports of participants’ autistic traits and adaptive functioning, participants’ self-report and projective tests for self-assertiveness and psycho-social functioning, and parent reports on parental reflective functioning and perceived school support. Common process factors and their effects on outcomes will also be explored. Cost-effectiveness will consider both societal and healthcare perspectives.

**Ethics and dissemination:** The current study protocol has been reviewed and ethics approval has been obtained from the Ethical Board Committee at the Third Affiliated Hospital of Sun Yat-sen University (II2023-119-01). The trial was pre-registered in Chinese Clinical Trials (ChiCTR2300072136) on 2023 June 05. The results of this trial will be actively disseminated through peer-reviewed publications and conference presentations. Any identifiable personal information will be anonymised to protect confidentiality.

**Protocol version:** 2.0, 2024 July 31

**Strengths and limitations of this study:**

- Comprehensive assessment of diverse outcomes via multiple informants.
- Systematic cultural adaptation using the Integrative Cultural Adaptation Framework (ICAF)
- ensuring the appropriateness of the Chinese version of KONTAKT™.
- Collect data on readiness for change, providing insights into “help-seeker” effects.
- Limited generalisability due to a single study site, location-specific inclusion, and absence of an active comparator.
- Given the absence of previous research examining the efficacy of SSGT programs in autistic children in the Chinese Mainland, this study is appropriately a feasibility study. As such, only the preliminary efficacy of KONTAKT™ with this population in a small sample will be examined. The small sample size will limit the power of the study in examining the effect of moderators on the study outcomes.

## Introduction

### Background and rationale

Autism^1^ is a neurodevelopmental condition characterised by persistent challenges in reciprocal social interaction and communication [1]. Autistic children often struggle to comply with social expectations and norms of communication and interaction, impacting their functioning in everyday contexts [2]. The estimated prevalence of autism among children aged 6 to 12 in the Chinese Mainland is 0.7% (∼ 1 in 143 children) and nearly half of all autistic children attend mainstream schools [3]. While there is limited research investigating the experiences of these children, their families and teachers, there is a clear and urgent need for evidence-based interventions aiming to support the social competence and well-being of Chinese autistic children in their everyday lives.

Internationally, Social Skills Group Training (SSGT) programmes are demonstrating efficacy in supporting verbal and cognitively able school-aged autistic children in developing their social skills [4, 5, 6]. SSGTs facilitate socio-emotional development and promote well-being in autistic children by explicitly and implicitly supporting autistic children in interacting with peers and participating in society more broadly [7]. SSGTs also leverage the benefits of learning in a group, with opportunities for peer learning and support. Multiple meta-analyses of randomised controlled trials (RCTs) evaluating the efficacy of these programmes in autistic youth report moderate to large effect sizes in achieving personally meaningful social goals, enhancing social skills, social skills knowledge, social participation, and social functioning [4, 8, 9, 10].

While there has been considerable investigation on the proximal intervention outcomes of SSGT programmes in autistic children and adolescents and post-test and short-term follow-up periods, there has been limited focus on longer-term outcomes and the generalisation of learnt skills to everyday contexts such as schools. Further, with few exceptions, the outcomes of these programmes have been largely assessed by parents. This is of particular concern given that most study designs have not employed single-blinded observation of outcome measures [4, 8]. Results from previous meta-analyses [4, 8, 11, 12] infer that improvements observed in parent and clinical ratings were primarily related to increased social skills knowledge or specific task-related skills, rather than assessments of the generalisation of skills to everyday contexts [8].

SSGT programmes have also been criticised for failing to consider the view of autistic youth themselves in the design and delivery of the programmes [13]. While capturing and accounting for the views of autistic young people may require effort, given their communication challenges [14], increasingly autistic advocates and the Autistic community are calling for researchers and clinicians to consider and incorporate their views in clinical interventions [15]. It is also likely that co-producing SSGT with individuals with lived experience will enhance the efficacy of these programmes in relation to the intended target group [16]. Further, given the inherent challenges experienced by autistic children in understanding social interactions and emotional content, questions have been raised regarding the validity and accuracy of self-report questionnaires aiming to evaluate the impact of SSGT programmes on these children [17]. Despite these cautions, findings from a recent meta-analysis suggest that the emotional self-awareness of school-aged autistic children is comparable to their non-autistic peers [18]. It seems plausible that approaches tailored to the communication needs and preferences of autistic children will be more successful in gathering their perspectives [19]. Further, while unusual in SSGT or autism research context, validated non-verbal psychometric assessments, such as the Draw-A-Person Test [DAPT] [20, 21], may be a worthwhile tool for understanding children’s psycho-social functioning [22, 23, 24].

Methodological limitations of previous SSGT efficacy studies include a failure to consider common process factors [25]. Process factors are elements that are inherently present in receiving and delivering a health intervention, that unfold progressively throughout its delivery and include participants’ expectations, willingness to participate, and the alliance between participants and facilitators. While a significant proportion of SSGT studies have reported programme adherence and satisfaction [8], the data obtained by these studies were largely descriptive and failed to systematically examine the impact of process factors in relation to intervention outcomes [25].

The KONTAKT™ programme is a well-established SSGT designed for autistic children and adolescents with and without cooccurring conditions [26]. Originating in Germany [27], it has now been further developed, standardised, culturally adapted, and evaluated in Sweden [28, 29] and Australia [30]. Three session-length variants (24-session, 16-session, and 12-session) have been standardised and evaluated through RCTs, demonstrating efficacy and effectiveness in improving participants’ social functioning, adaptive functioning, emotional well-being, emotion recognition and regulation, as well as accomplishment of personally-meaningful social goals [28, 29, 30]. Findings across these studies suggest that KONTAKT™ may be particularly beneficial to autistic teenagers and girls. Moreover, both quantitative and qualitative data support the conclusion that KONTAKT™ is effective, safe, and well-received by autistic youth [28, 29, 30, 31, 32].

To our knowledge, while a standardised SSGT programme PEERS^®^ has been tailored for use with Chinese autistic adolescents [33], there is no culturally standardised SSGT programme specifically tailored toward the needs of autistic children years in the Chinese Mainland (See **Supplementary Table 1 [online supplemental file 1]** for a comparison between the KONTAKT™ CHILD and the Mandarin PEERS^®^; see also Afsharnejad *et al.* (2023) *pp.*1286-1287 & Table 4 [6]). With its diverse, individualised, and naturalistic session design [26], it is anticipated that KONTAKT™ will effectively support the social functioning and well-being of Chinese autistic children, provided the context of Chinese culture and its associated social norms are incorporated [34].

Cultural adaptation involves systematically modifying an evidence-based intervention to align with the target population’s needs, values, languages, conditions, and customs [35, 36, 37]. However, only a few cultural adaptation studies on SSGT have well-documented and theoretically based methodologies. Moreover, previous SSGT efficacy studies generally lacked input from the target population, despite language translation and the inclusion of culturally relevant content [6].

Ignoring cultural differences can significantly hamper the effectiveness of evidence-based SSGTs for autistic individuals [34]. Given the core characteristics of autism involve social interaction and communication, social-ecological differences across cultures will undoubtedly influence interpretations of “functioning” and “disorder” in autism [38]. Cultural factors also impact established expectations and interactions between families and service providers, and between service providers in various sectors including healthcare, education, and the broader community [39, 40, 41].

For example, Chinese parents are unlikely to disclose their child’s autism diagnosis to school, reflecting the limited collaboration between family-school, prevailing autism stereotypes and fears regarding stigma [42]. Moreover, Chinese autistic children are commonly not informed of their autism diagnosis, with the majority of parents choosing not to disclose the diagnosis to them [43]. There are also clear differences in the prevailing service delivery paradigms relevant to supporting autistic children in the Chinese Mainland compared to many Western countries. Service delivery in the Chinese Mainland is significantly impacted by a large population, and limited health professionals and public health services [44, 45]. Unlike many Western countries, where funding and child health services for autistic children are commonly provided by governmental departments, the Chinese Mainland relies primarily on parents to cover these costs [46].

While there are various cultural adaptation frameworks guiding the alignment of health interventions with the needs of a target population [35, 36, 47, 48], despite some methodological distinctions, they share many similarities. In aligning the KONTAKT™ SSGT programme with the needs of autistic children living in the Chinese Mainland, this study adopts the integrative cultural adaptation framework (ICAF) [35]. The ICAF involves five stages: information gathering, preliminary design, preliminary efficacy testing, refinement, and final trial. This study focuses on the first four stages of the ICAF, in preparing for final evaluation trial (the last stage).

### Objectives and hypotheses

The overarching purpose of the present study is to assess the feasibility and cross-cultural validity of the adapted 16-session version of the KONTAKT™ CHILD programme for Chinese autistic children in preparation for a larger RCT trial. The specific objectives are:

1. Evaluating the integration of KONTAKT™ CHILD programme into a tertiary hospital’s routine activity;
2. Assessing the acceptability and feasibility of the data collection framework;
3. Examining the preliminary efficacy, quantitative and qualitative effects of KONTAKT™ to inform a future, definitive RCT;
4. Exploring the impact of common process factors on intervention outcomes; and,
5. Examining the cost-effectiveness of KONTAKT™ CHILD compared to standard care.

Our primary hypothesis is that the Chinese 16-session KONTAKT™ CHILD programme will be feasible and culturally appropriate for Chinese autistic children with minor revisions and can be successfully implemented in a tertiary clinic. We also hypothesise that participants in the immediate training group (ITG), in comparison to the delayed training group (DTG) [wait-list], will experience greater improvements in outcomes from Time 1 to Time 3 (waiting period of DTG). We expect that participants and their parents with higher readiness for change and a stronger alliance in the ITG will benefit more from KONTAKT™ CHILD. Moreover, we anticipate that the training effects in the ITG will persist at follow-up, and significant improvements in children’s outcomes will occur in the DTG from Time 3 to Time 5 (training endpoint of the DTG). For cost-effective analysis, we hypothesise that the additional costs for KONTAKT™ will be offset by participants’ improvement in health-related quality of life and out-of-pocket savings in the use of other mental health and psychiatric services.

## Trial Design

This study was approved by the Ethical Board Committee at the Third Affiliated Hospital of Sun Yat-sen University (II2023-119-01) and preregistered in Chinese Clinical Trials (ChiCTR2300072136).

Any modification to this protocol will be reported to the ethical board and updated in Chinese Clinical Trials. Following Thabane and Lancaster’s guidelines in 2019 [49], this Chinese KONTAKT™ CHILD protocol is planned and reported following the Standard and Protocol Items: Recommendations for Interventional Trials (SPIRT) [50]. The findings of this study will be reported adherent to the Consolidated Standards of Reporting Trials for Social and Psychological Interventions (CONSORT-SPI) [51].

The Chinese KONTAKT™ CHILD feasibility study has a prospective, wait-list controlled, single-site, stratified, randomised design. Blinding will apply to data collection, outcome adjudicators, and data analysts. All electronic files, including coding videos and scanned drawings, will be named without identifying personal information and group allocation or time-point information. Three major assessment time-points (ITG: Pre-training [T1], Post-training [T3], and Follow-Up [T5]; DTG: Pre-training [T1], Pre-training [T3], Post-training [T5]), with additional mid-point cost and processing factors data collection for each group, are included. Additional mid-point cost data collection follows the two-month follow-up intervals recommended by the Treatment Inventory of Costs in Patients with Psychiatric Disorders - Child version (TiC-PC) manual [52]. Measurements will be taken during face-to-face meetings, with the exception of data from teachers and the prescreening process. Further, this study will employ a mixed-methods design, with both between-group and within-participant comparisons, to explore the feasibility, acceptability of implementing a large-scale RCT, and the preliminary efficacy of the Chinese KONTAKT™ CHILD programme. Randomisation is performed at a 1:1 allocation ratio, with variable block length and stratification by sex. Participant recruitment and training are ongoing between 2023 June 5 and 2025 March 1.

## Methods: participants, interventions, and outcomes

### Study setting

All in-person assessments and interventions, except for the excursion session, will take place at the outpatient department of the Child Development and Behaviour Centre (CDBC) of the tertiary academic hospital, the Third-affiliated Hospital of Sun Yat-sen University, in Guangzhou, China. The CDBC is one of China’s first established and leading developmental behavioural paediatric units, with over 20 years of experience in diagnosing and promoting the well-being of autistic individuals and their families. The CDBC provides over 10,000 clinic appointments annually to neurodivergent children and their families across China seeking diagnostic and intervention services.

Online assessments will be designed using the SurveyStar online survey platform (https://wjx.cn/). The SurveyStar is one of the most popular electronic survey platforms in Chinese contexts, with its services covering more than three million enterprises and 90% of universities in China.

Due to limited email use in the Chinese Mainland, WeChat^©^, the dominant instant messaging and media app, will be the primary channel for participant recruitment, communication with families, and online survey distribution. Additionally, the SurveyStar conveniently integrates with WeChat^©^ through a Mini Programme [53], allowing WeChat^©^ users to seamlessly complete questionnaires directly on the platform.

The KONTAKT™ certified trainers are experienced CBDC clinicians (clinical psychologists, developmental behavioural paediatricians, and therapists) working with Chinese autistic children (with 3 to 10 years of working experience). They have completed standardised online KONTAKT™ methodological training provided by experienced trainers from the Center of Neurodevelopmental Disorders at Karolinska Institutet (KIND), including self-directed training and video-conference seminars, followed by a 13-session enhanced in-person training. The enhanced training course mainly included trainers role-playing the child sessions, and more in-depth discussion on the principles of KONTAKT™.

All KONTAKT™ CHILD sessions, except for the excursion session, will be delivered in a spacious, well-lit room at CDBC, with participants and trainers sitting in a circle. As suggested in the KONTAKT™ manual, delivery of the programme content will be supported by a flipchart, utilised to record notes during each session, and a board displaying the group rules, session agenda, and points assigned to participants in recognition of their participation in the sessions. The excursion session (session 9) will be to a café, a bubble tea shop, or a convenience store, accessible from CDBC. Parent sessions will take place in a separate room, with parents sitting together in a circle without their children present. A projector will be utilised to present information on the principles and the content of KONTAKT™, and information pertaining to their role, such as completing assignments and providing constructive feedback, in supporting their children during the intervention period.

### Recruitment

**Figure 1** displays the flow of participants throughout the study. Thirty-six autistic children will be recruited through the outpatient department of the CDBC in two rounds of recruitment. Multiple recruitment avenues will be utilised including word-of-mouth, referrals from clinicians, posters displaying information about the study at the CDBC clinic, and flyers published on the WeChat^©^ account of CDBC (https://www.wechat.com/).

**Figure 1.**
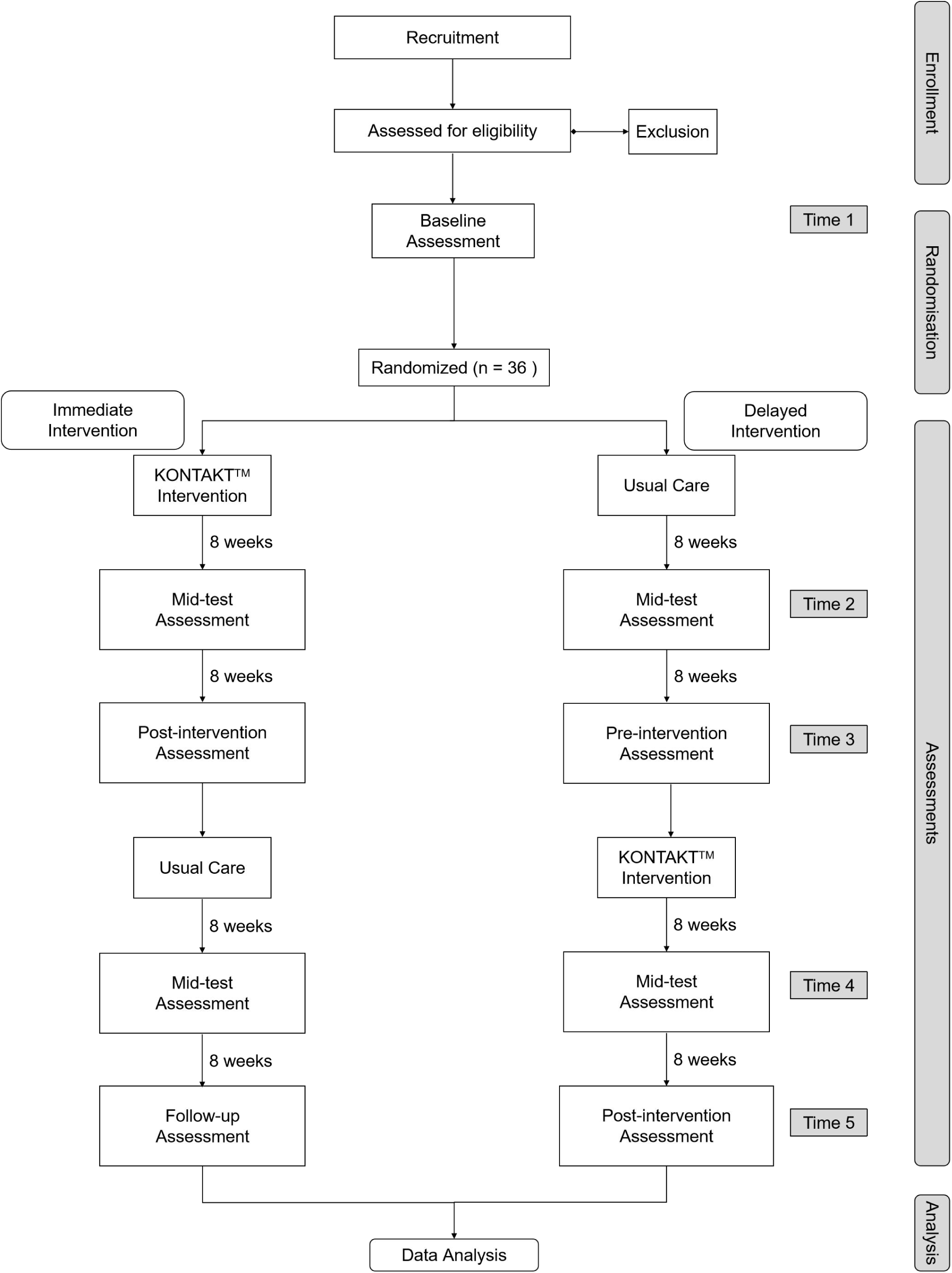
**Chinese KONTAKT™ CHILD feasibility study flowchart.**

Parents will express their interest in the study by completing a brief online registration form (taking approximately 10-15 minutes to complete) obtaining their contact information, their child/children’s demographic information, diagnostic statuses, previous evaluations on the Wechsler Intelligence Scale for Children-IV Chinese version (WISC-IV) [54], the Autism Diagnostic Observation Schedule-Generic (ADOS-G) [55], and screening for problem behaviours which may impact on their ability to engage positively in a group setting using the Childhood Behavior Checklist-parent version (CBCL) [56]. Obtaining parents’ expression of interest in this way enables estimation of the demand for KONTAKT™ from families engaging with CDBC.

Following the submission of their expression of interest, a research coordinator will assess candidates’ eligibility to participate in the study. Parents of children meeting the inclusion criteria for the study will subsequently participate in a 10-20 minute prescreening phone interview, discussing with the research team their child’s school lives (e.g. how their child is getting along with their classmates and teachers), academic performances, and special needs, and addressing any study-related questions. This brief interview with parents will also ascertain potential participants’ suitability to cope with the cognitive and verbal demands of KONTAKT™.

If fulfilling the prescreening criteria, candidate participants and their parents will attend CDBC for a 50-minute face-to-face intake interview with the research coordinator. During the intake interview, both children and parents will participate in a 20-minute individual interview with the research coordinator. Families meeting the inclusion criteria of the study will be provided with detailed oral and written information about their participation in the study, including the requirements. Written informed consent will be obtained from parents and children.

### Eligibility criteria

#### Inclusion criteria

1. Aged 8-12 years;
2. A clinical diagnosis of autism (confirmed by ADOS-G, module 3);
3. A Full-scale IQ > 70 on the WISC-IV; and,
4. Resided in Guangzhou city for the past 12 months.

#### Exclusion criteria

1. Significant physical impairment or severe metabolic disease(s);
2. Significant rule-breaking or aggressive behaviours within the last six months, as confirmed by CBCL;
3. Low intrinsic motivation to participate as confirmed by the intake interview;
4. Significant difficulties with Chinese characters reading comprehension or Mandarin expression and comprehension; and,
5. Prior or current behaviour or psychiatric conditions that could disrupt participation or necessitate other treatment(s).

Participants with common co-occurring neurodevelopmental or psychiatric conditions in autistic individuals, such as attention-deficiency hyperactivity disorder, depression, anxiety, and tic disorder, are eligible if these conditions do not significantly interfere with participation. Participants will continue with their prescribed pharmacological treatments, psychological therapies, or other interventions while participating in KONTAKT™ CHILD. Written informed consent will be obtained from participants and parents after a thorough explanation of the study during the intake interview.

### Intervention

#### KONTAKT™ CHILD Intervention

Prior to the development of this study protocol, a preliminary adaptation of KONTAKT™ (The first step of the ICAF - Information gathering) has been completed. The KONTAKT™ programme includes workbooks for trainers, children, and parents, supporting intervention fidelity. In adapting the 16-session variant of KONTAKT™ from the Australian English version to Mandarin Chinese, a rigorous process was employed. Firstly, an initial translation was made and verified for accuracy by bi-lingual authors (LU, ZH, and EW). Then, a working group of KONTAKT™ Chinese trainers, who are also CDBC clinicians, provided feedback on the initial programme materials. This step resulted in significant modifications to the discussion topics, particularly regarding those addressing autism diagnosis, making them more general and implicit (most notably in session 3). In addition, to encourage parents to discuss the autism diagnosis with their child, information relating to “how to talk about the diagnosis with your child”, was added to the first parent session. This adaptation was made given that research has highlighted that over 60% of Chinese autistic children are unaware of their autism diagnosis, with 40% of parents of Chinese autistic children preferring not to disclose an autism diagnosis to their children [43].

The current adaptation of the KONTAKT™ programme also emphasises enhanced parent education and support. Parents will receive information and resources to support their children with the KONTAKT™ homework activities. This includes parents role-playing parent-child interactions during homework completion contexts and demonstration videos. Throughout the programme, KONTAKT™ trainers will also provide session-by-session feedback, interim and final report to parents on their child’s progress. Additonally, they will offer ongoing coaching and encouragement to parents. This enhanced parent support directly addresses a need identified in previous research [33] - Chinese parents expressed a desire for additional coaching skills education to facilitate their autistic children’s participation in SSGTs.

Moreover, members of the Chinese research team further modified and added questions, games, activities, and scenarios aligned with Chinese cultural norms. A Chinese name was also created for KONTAKT™, emphasizing the training’s focus on meaningful socialisation (交得益 ™). Finally, a family with an autistic adolescent proofread the manual and made edits to enhance language fluency and readability, and the inclusion of culturally appropriate terminology. Additionally, a logo for the Chinese version of KONTAKT™ was designed and utilised on customised stationary and course materials, to foster a sense of group cohesion among participants.

KONTAKT™ integrates Cognitive Behaviour Therapy principles and learning principles tailored towards the needs of autistic youth (see **Figure 2**) [26]. These include principles of psychoeducation, observational learning, modelling, behavioural activation, homework, computerised cognitive training, opportunities for rehearsal, a structured agenda, and parental involvement. KONTAKT™ employs mandatory, recurring, and variable activities to support participants in achieving their personally meaningful social goals. Sessions aim to enhance understanding of social norms, develop adaptive socio-emotional skills, and promote practical applications. The first 11 sessions of KONTAKT™ consistently present weekly themes and assignments, while later sessions are led by the participants and tailored towards topics of interest to each group. In this way, KONTAKT™ accommodates not only for cultural context but also for the subtle differences and preferences of each KONTAKT™ group.

**Figure 2.**
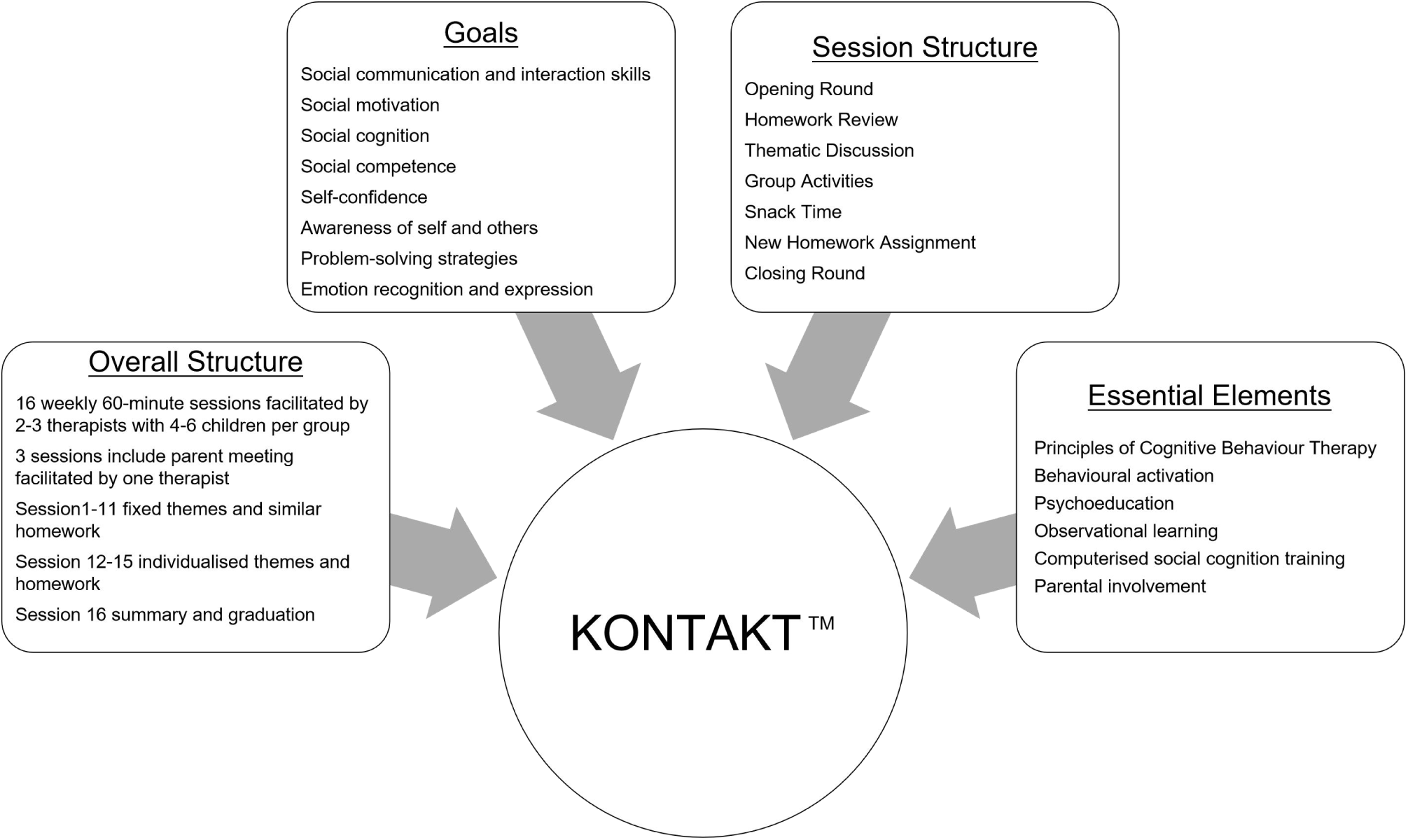
Elements, structures, and goals of KONTAKT™. [7, 26].

A group of 4-6 participants will attend 16 consecutive sessions of KONTAKT™ led by 2 to 3 trainers on weekends, with each session lasting approximately 60 minutes. Three parent sessions will take place at the beginning, middle, and end of the training, offering parents individual feedback on their child’s progress, presenting educational information on autism and approaches to discussing an autism diagnosis with their child, and encouraging parents in socially coaching their children. During the parent sessions opportunities will be provided encouraging parents to ask questions, provide feedback, and engage with other parents of autistic children. Parents will be encouraged to assist their children with homework, with trainers providing feedback to parents after each session. Refer to **Supplementary Table 2** (**online supplemental file 1**) for the structure and content of the Chinese KONTAKT™ CHILD version.

To ensure fidelity to the KONTAKT™ training protocol, trainers will complete session-by-session fidelity checklists and receive supervision from certified KONTAKT™ trainers.

### Description of the rationale for adopting the Chinese 16-session KONTAKT™

While KONTAKT™ has been previously standardised and published in 12 and 24 session variants, the reasons underpinning the choice to tailor the 16-session variant to the Chinese Mainland were three-fold. Firstly, previous research found that the 12-session variant did not significantly outperform standard care in Sweden in children [28], and the 24-session variant had a higher attrition rate than the 16-session variant and 12-sesion variant (26% vs 11%/14%) [28, 29, 30]. Moreover, aligning with the Chinese academic year structure, which consists of two terms with approximately five months each, the 16-session version is more practical. Lastly, evaluating the 16-seesion variant will result in the DTG waiting for a 4 to 5 month period, which would be extended by evaluating the 24-session variant and negatively impact on adherence and the timeliness of access to the intervention [57].

### Explanation for the choice of comparators

The waiting period of the DTG is limited to 4-5 months, roughly equivalent to one round of KONTAKT™ training. While an active comparator in an RCT is typically the most robust, due to limited clinical resources in the Chinese Mainland, a wait-list control group is considered more practical. Moreover, given that “nonspecific” care is common in the Chinese Mainland, a wait-list control group allows for assessing the additional impact of KONTAKT™ CHILD in addition to standard care.

### Intervention adherence

We will use a participant attendance sheet to track compliance, aiming for at least 80% attendance (13 out of 16 sessions) as acceptable. Homework completion will also be recorded, with a target of over 60% considered tolerable.

We will further adopt several approaches as recommended by Robiner to enhance the wait-list group’s adherence [58]:

1. Provide detailed study information to families during intake interviews;
2. Emphasise to families the importance of adherence;
3. Establish a collaborative working relationship with families during intake interviews to develop trust and rapport;
4. Offer face-to-face assessments for regular clinical visits during the waiting periods;
5. Limit the waiting time for the DTG to 4-5 months with a single-round intervention; and,
6. Schedule appointments at times convenient to families, for example on weekends.

### Randomisation

After the initial visit to the clinic, participants will be randomised to either ITG or DTG. Randomisation will be conducted by a third party not involved in the Chinese KONTAKT™ CHILD trial using computer-generated sequentially random numbers. The permuted block numbers 4 to 6 will be used to restrict randomisation within the strata (birth-assigned sex). Each participant will be assigned a unique code.

### Assessments/measures

Outcomes will be assessed from the perspective of multiple informants (children, parents, teachers, clinical ratings), via multiple data collection methods (questionnaires, interviews, observations, projective tests). The data collection timeline is presented in **Supplementary Table 2 (online supplemental file 2)**.

All online questionnaire fields are mandatory, requiring completion to progress, eliminating the expectation of missing data. QR codes will be used for distributing the registration and teacher questionnaires, completed via the WeChat^©^ app. All teacher questionnaires will be assigned a unique participant identification number.

*Primary outcome measures* will inform the feasibility of conducting a full RCT. Feasibility outcomes include acceptability and appropriateness of intervention, and negative effects assessed via quantitative data (fidelity checklists, recruitment rates, attendance rates, homework completion rates, Group Session Rating Scale [59], Theoretical Framework of Acceptability Questionnaire [60], and Negative Effects Questionnaire [61]) and qualitative data from focus groups with both participants and providers conducted post-intervention (ITG: post-training [T3] and follow-up [T5]; DTG: post-training [T5]). The relationship between the focus areas of feasibility studies, as outlined by Bowen *et al.* [62] and those addressed in this study is summarized in **Table 1**. **Table 2** provides a detailed description of the quantitative feasibility parameters and criteria for success.

**Table 1.**
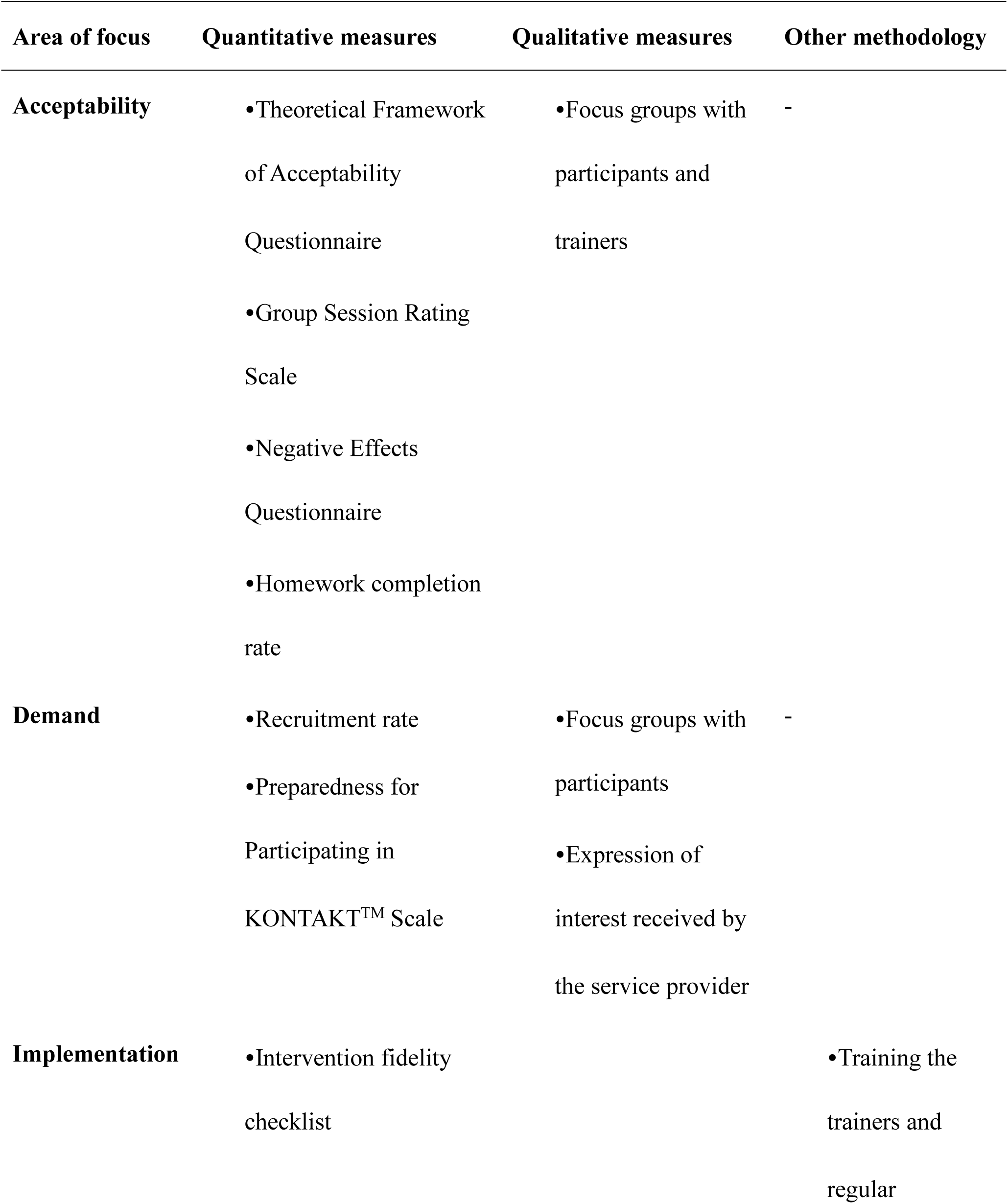
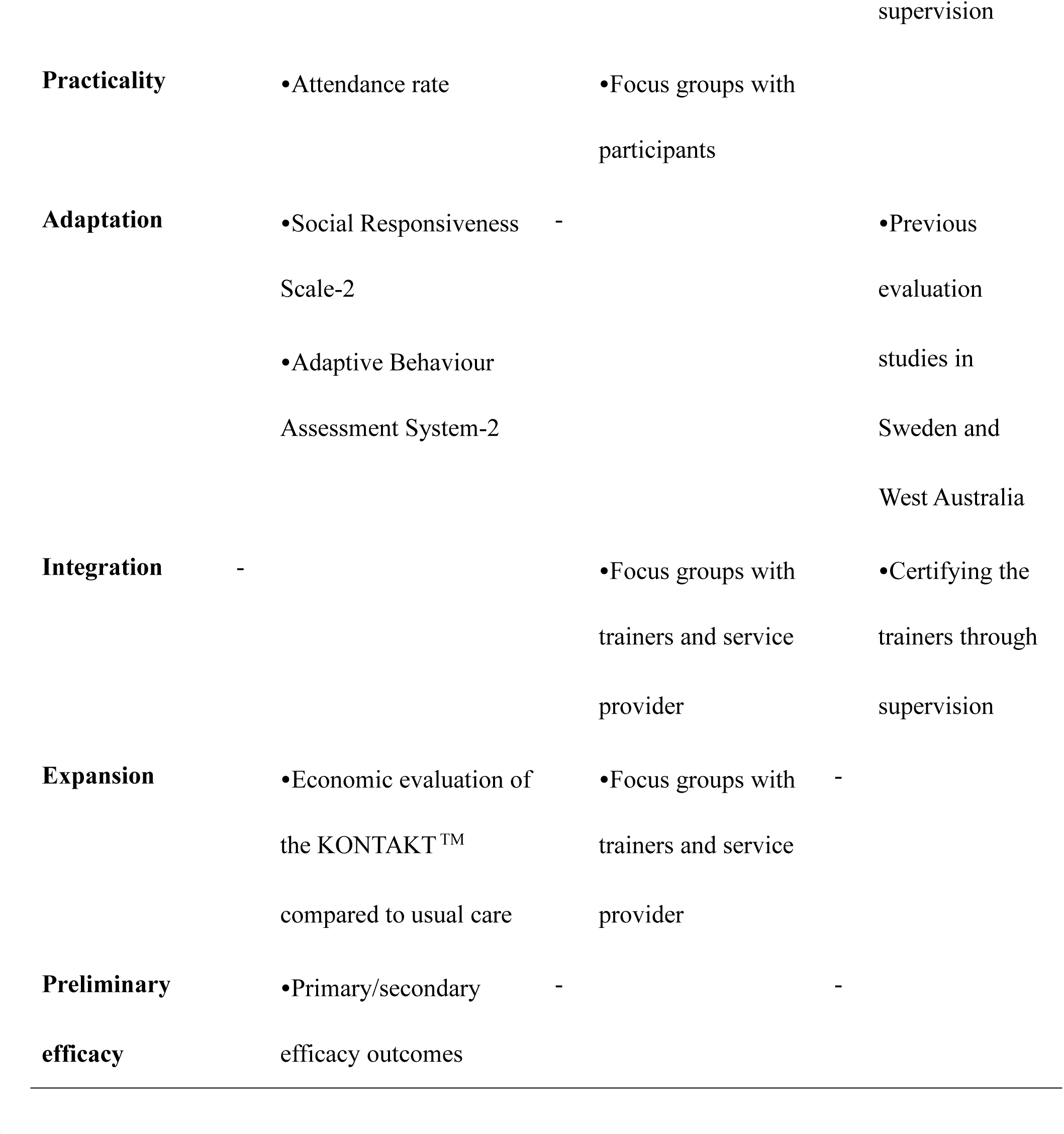
Feasibility methodology based on the focus areas outlined by Bowen et al. (2009) [62]

**Table 2.**
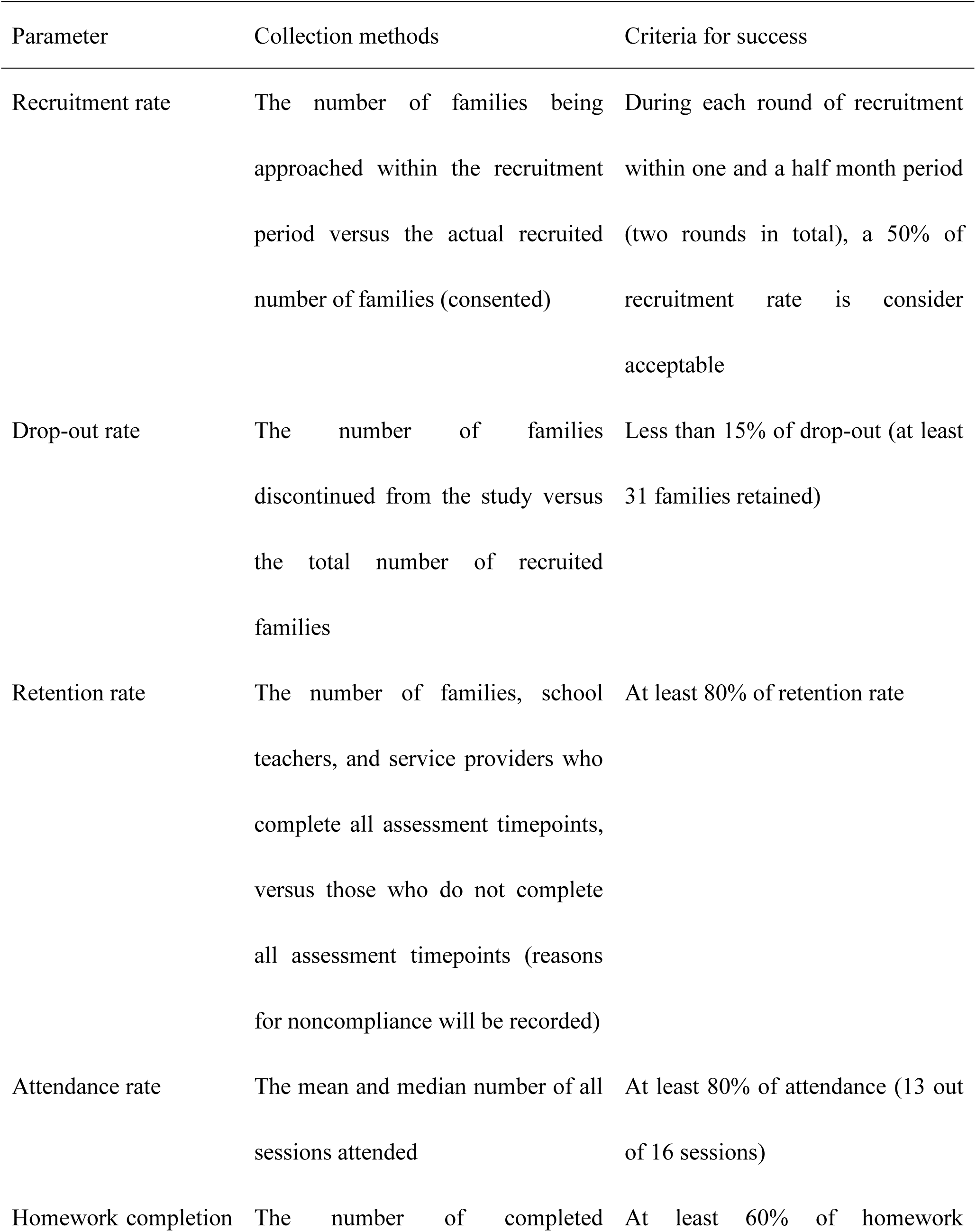
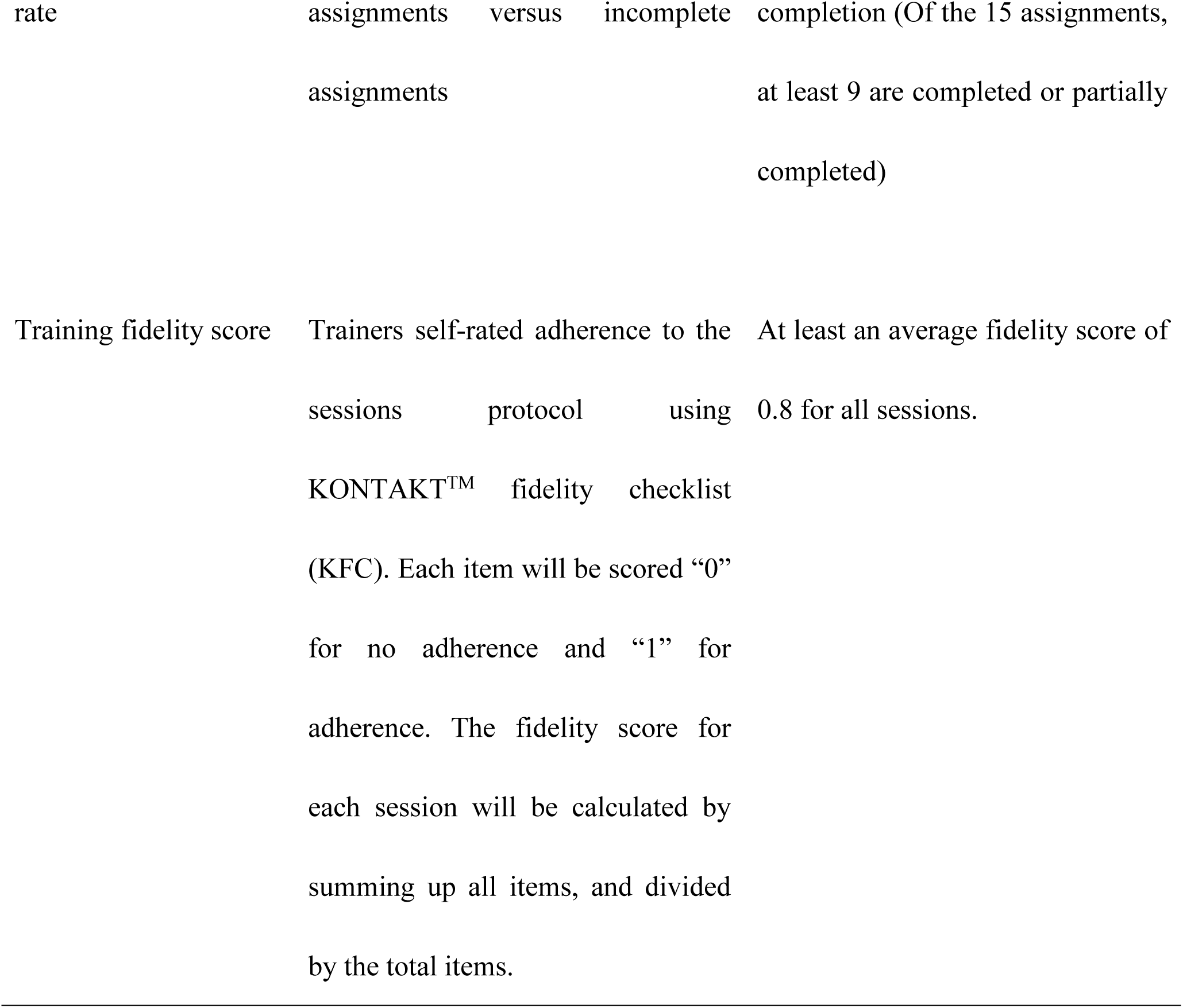
Feasibility parameters and criteria for success.

*Secondary outcomes* will focus on the preliminary efficacy of the Chinese 16-session KONTAKT™, which will inform the future full-scale RCT. Outcomes include the following measures:

The primary efficacy outcome of the KONTAKT™ CHILD study is improvement in social communication and interaction skills. This is assessed using the Contextual Assessment of Social Skills (CASS), an ecologically valid observational measure [63]. The CASS involves two videotaped role-play assessments where participants engage in reciprocal conversation and interaction. These assessments take place in contexts that are manipulated to be either interesting or boring. This manipulation is achieved by changing how the confederate responds and interacts with the participant. During each role-play, participants and an unknown adult confederate of an opposite birth-assigned sex pretend that they have just joined a new club and are waiting for the first session to begin. During this three-minute waiting period, participants and the adult confederate can talk to each other.

Secondary efficacy outcomes include autistic characteristics (Social Responsiveness Scale-2 [SRS-2] [64]), generalised social skills (Snack Time Assessment of Relationships and Social Skills [STARSS] [63, 65]), adaptive functioning (Adaptive Behaviour Assessment System-II [66, 67]), self-esteem and self-concept (Self-concept Scale Draw-A-Person [SCS-DAP] [68], Child Rosenberg Self-Esteem Scale [CRSES] [69]), cognitive development and psychosocial well-being (Koppitz Draw-A-Person Test [70, 71], Child Outcome Rating Scale [72, 73]), perceived school support school level [33], and parental reflective functioning [74]. *Common process factors measurements* include alliance (Therapeutic Alliance Scales for Children [75, 76]) and readiness for change (Preparedness for Participating in KONTAKT™ Scale [77]). *Cost-effectiveness outcomes* include healthcare consumption and productivity loss (the tailored version of the Treatment Inventory of Costs in Patients with Psychiatric Disorders - Child Version [78, 79]) and health-related quality of life (Child Health Utility 9D [80], EQ-5D-Y [81]).

For details of these measures, see **Online supplemental file 2**.

### Retention

Families will be requested to attend data collection appointments at a time convenient to them with reminders from research assistants. Paper-pencil questionnaires will be mailed to families unable to attend appointments in person and provided with a pre-paid envelope to return these via post. The reasons for participant withdrawal, adverse events, and other unintended effects will be documented and reported, and drop-outs will not be replaced.

### Data management

In the Chinese KONTAKT™ CHILD feasibility study, all paper data will be converted to electronic format. Trained research assistants will input the data within two weeks after each assessment. Both paper and electronic data will be stored in EpiData [82] with double-entry for data integrity. Checks for referenced range values, valid values, and regular data consistency will be applied. Non-numeric data, including open-ended questions and notes, will be entered as originally written.

Video and audio recordings will be labelled with unique identifiers, with details recorded in the EpiData repository. Paper documents will be securely stored in locked filing cabinets at CDBC. Electronic textual data will be password-protected. Video and audio files will have dual backups on encrypted hard disks. Only the principal investigator will have full data access.

Incremental data and recording files will be backed up daily.

### Sample size

The primary outcome of this present study is the CASS. Previous SSGT studies applying the CASS suggested a medium effect size (ES) of 0.42 [83]. Additionally, a rough average ES of 0.40, as measured by the SRS-2, is derived from the three prior RCT studies evaluating KONTAKT™ [28, 29, 30], and informs the expected ES for the future Chinese KONTAKT™ CHILD main trial. According to Whitehead *et al.* [84], a feasibility study with a sample size of 15 per treatment arm allows for detecting a medium ES (0.3∼0.7) with 90% power and two-sided 5% significance in the main trial. Accounting for a 15% attrition rate from previous KONTAKT™ evaluation studies, we require a sample size of *n* = 18 per arm (*N* = 36 in total).

### Statistical analysis

#### Statistical methods for primary and secondary outcomes

Feasibility measures will be reported descriptively. Additionally, qualitative data will be transcribed verbatim, de-identified, and then imported into NVivo 12 for theme-based analysis. If all or some feasibility parameters for success are not met (**Table 1** & **Table 2**), or if salient themes of unacceptability or unfeasibility emerged within the qualitative analysis, investigators will further examine the potential contributing factors, including participant baseline characteristics and session-composition characteristics. This analysis will inform necessary modifications to the KONTAKT™ CHILD training and its delivery in the Chinese Mainland.

For preliminary efficacy analysis, the primary efficacy outcome for this study is social-communication skills improvement as measured by the CASS, with the follow-up being the primary endpoint. The analysis will follow the intent-to-treat (ITT) principles [85], and all data from every participant will be analysed. Missing data will be handled based on established guidelines for each measure or CONSORT-SPI statement. No interim analysis will occur.

Data will be checked for normality and variance heterogeneity. Baseline demographics will be compared between ITG and DTG using t-tests or Wilcoxon rank test (continuous variables) and chi-square tests (categorical variables) to verify group comparability. Random mixed effects regression models, which have the advantage of handling missing data, will compare ITG and DTG for primary and secondary analyses, considering time (baseline, endpoint, follow-up), group (ITG, DTG), and their interaction variable (time × group). Participants’ ID will be regarded as a random effect accounting for intra-individual correlations. Intervention effects will be reported as least-square mean differences. ES will be reported by dividing the groups’ mean difference (baseline to endpoint and baseline to follow-up) by the pooled standard deviations, with positive values favouring the intervention. Potential moderators, including age, sex, IQ, co-occurring conditions and the common process factors at baseline between participants with high training responses (HR, defined as ≥ 30% improvement on CASS scores at follow-up), medium training responses (MR, defined as 10-30% improvement), and low training responses (LR, defined as ≤10% improvement or worsened) will be compared using either oneway ANOVA (continuous variables) or Chi-square tests (categorical variables) [31, 32, 33]. Post-hoc Dunn test or Chi-square test will be conducted if the ANOVA tests or multiple Chi-square tests demonstrate significant group differences. ES will be presented as either Hedge’s *g* or Cramer’s *V*.

#### Health economic analyses analysis

Cost-effectiveness analysis (CEA) will follow an ITT basis, considering societal and healthcare perspectives. Cost differences will be statistically tested, while resource usage differences will not be compared to prevent false positives. Quality-adjusted life year (QALY) will be calculated from EQ-5D-Y and CHU9D scores using validated algorithms for Chinese children and adolescents [86, 87]. Given potential non-normality in cost data, a non-parametric random forest permutation model will handle missing data. The incremental cost-effectiveness ratio (ICER) will be calculated as Δcost/ΔQALYs. Cost differences between ITG and DTG at different time points will be analysed via linear regression with bootstrapping for 5,000 repetitions to generate non-parametric confidence intervals (See **online supplemental file 3** for CEA details).

### Community and public involvement

The first author, LU, is self-identified as autistic. During the adaptation stage, the training manual for KONTAKT™ was proofread by a family with an autistic adolescent. To assess training burden and programme acceptability, participants will provide feedback through a combination of questionnaires and focus group interviews (mixed methods). Advocating for outcomes focused on well-being and mental health, autistic people continue to emphasise the importance of listening to their voices. Thus, this trial will also capture both quantitative and qualitative data examining participants’ experiences of receiving KONTAKT™ CHILD and the impact of the programme on children’s well-being. We will further engage the autism community, involving both parents with autistic children and clinicians, in recruiting potential participants. Finally, participants will be invited to share their lived experiences of participating in KONTAKT™ with the autism community through community events and social media in China when the trial has ended.

## Discussion

The study commenced recruitment on 2023 June 5. This article presents the design of the Chinese KONTAKT™ CHILD trial, a feasibility wait-list RCT featuring the Chinese 16-session KONTAKT™ variant for autistic children. This design incorporates multiple data sources and diverse outcome assessment methods, facilitating a comprehensive exploration of the feasibility of this programme from different perspectives. Furthermore, a systematic cultural adaptation process ensures the suitability, acceptability, and comprehensiveness of the Chinese KONTAKT™ version. Adhering to the ICAF framework, this study will guide the refinement of the Chinese KONTAKT™ version and future larger-scale evaluations, addressing the question of whether SSGT can be effectively implemented for Chinese school-aged autistic children.

While there are several strengths in the study design mentioned earlier, it’s important to acknowledge some limitations. Firstly, conducted in Guangzhou, one of China’s most advanced cities, the study’s single-site nature may limit the generalisability of the findings to other clinical settings in the Chinese Mainland, especially to regions with more limited resources. Additionally, certain measures planned for use in the study, such as the CRSES and the SCS-DAP, may have uncertain psychometric properties due to the lack of Chinese population data. However, this feasibility study aims to provide empirical data on the psychometrics of these measures with Chinese autistic children. Another limitation is the absence of an active comparator, as the wait-list control could potentially overestimate treatment effects [88]. Nevertheless, this aligns with the prevailing situation in China. Furthermore, the study will collect data on parents’ and children’s readiness for change, allowing for an exploration of the “help-seeker” effects, following the recommendation by Cunningham and colleagues in 2013 [88].

As mentioned above, the final stage of the ICAF involves running a full-scale RCT trial to evaluate the efficacy of the Chinese version of KONTAKT™. Therefore, upon completing this feasibility study, the investigators will integrate all quantitative and qualitative results to decide whether a full-scale RCT trial will progress or redesigning another KONTAKT™ feasibility study will be needed. The following aspects will be considered: (1) participants’ and parents’ acceptability and adherence rates, (2) participants’ and parents’ feedback on demand and training burden for KONTAKT™, (3) trainers’ feedback on implementing KONTAKT™ in Chinese clinical setting, (4) trainers’ fidelity on implementing KONTAKT™, (5) the feasibility results of the processes for data collecting, and (6) considering the potential capacity of the intervention setting (human resources, materials, and equipment) on implementing a full-scale RCT regarding the sample size calculation for the primary outcome.

## Plans for dissemination of finding

The findings of the Chinese KONTAKT™ CHILD trial will be submitted to peer-reviewed journals for consideration. Abstracts will also be submitted to both local and international conferences, and community-based lay abstracts will be written and disseminated to provide feedback to autism stakeholders in China.

## Data Availability

The ethical consent and funding documents have been reviewed by the review board of the Chinese Clinical Trial Registry and are available for review by the Editorial Office. Anonymised data related to this study will be available from the corresponding author upon reasonable request after study completion and publication of the study. The final dataset generated during this study will upload to the Research Data Deposit of Sun Yat-sen University. Trial results will be reported and published in scientific journals.

## Authors’ contributions

LU, ZH, ZX, SB, and SG undertook the conceptualisation and design of the study. LU, ZH, EW translated the manuals and workbooks. LU, ZH, SB, SG, and ZX selected the measures used. EW, FJ, YP, HX, ZH, LU, and ZX improved the content of the manual. FJ, YP, ZH, and ZX assist with administration of groups and offer clinical support. LU drafted the manuscript, with inputs from ZH, SB, SG, and ZX. All authors contributed to, read and approved the final manuscript.

## Ethics approval

This study has been approved by the Ethical Board Committee at the Third Affiliated Hospital of Sun Yat-sen University (II2023-119-01), and pre-registered in Chinese Clinical Trials (ChiCTR2300072136).

## Acknowledgments

We acknowledge the generous support from the Swedish team at Karolinska Institutet/Stockholm Health Services and the Australian team at Curtin University. They facilitated the training, provided precious experiences and materials for the CDBC KONTAKT™ trainers. We thanks the CDBC KONTAKT™ team for facilitating the process of this trial. We sincerely thank the autistic individual, Yihan Liu, and her families for validating the translation and contents of the Chinese KONTKAT™ manual. Additionally, we appreciate Yan Li for designing the tools for the KONTAKT™ CHILD programmes.

## Footnotes

1 Herein, identity-first language is used since this is the endorsed terminology among autistic advocates (Kenny, 2016).

