## Supplementary material for "Feasibility and cross-cultural validation of an adapted social skills group training programme (KONTAKT™ CHILD) for Chinese autistic children: a wait-list RCT protocol": online supplemental file 1

### Online Supplemental File 1: Structure and Content of the Chinese version of KONTAKT™

Supplementary Table 1. Comparison between the KONTAKT™ CHILD and Mandarin PEERS® for autistic children and adolescents.

| **Category** | | **KONTAKT™ CHILD** | **Mandarin PEERS®**[1] |
|---|---|---|---|
| Participant characteristics | Age range (in years) | 8-12 | 12-17 |
| | Intelligence quotient (IQ) | Full-scale IQ over 70 | Full-scale as well as verbal IQ over 70 |
| | Adequate Chinese comprehension | Required | Required |
| | Self-reported intrinsic motivation to participate | Required | Required |
| | No significant rule-breaking or self-harm behaviours | Required | Required |
| General group and session characteristics | Number of participant sessions | 16 | 14 |
| | Number of parent sessions | 3 | 14 |
| | Group size | 4-8 | 6-10 |
| | Per session length | 1 hour | 1.5 hour |
| | Number of therapists per training group | 2-3 group leaders | 3 group leaders for the adolescent session and 1 for the parent session |
| Session structures and parent involvement | Participant session structure | Structured, with various sections including group activities and snack times | Structured, with the main focus being the didactic section |
| | Participant-led sessions, individualised topics, and individually tailored assignments | Yes | No |
| | Excursion session for skill generalisation | Yes | No |
| | Parent session structure | Focus on the overall progress of participants and group; parental support | Focus on homework review and parental coaching techniques |
| | Parent involvement | Less emphasised, parents are encouraged to support their children throughout the programme | More emphasised, and successful parent coaching is considered as elemental in promoting positive training outcomes |
| | Teaching methods | Scaffold type teaching | Didactic teaching |
| Overall goals and guiding principle | Overall goals | Individualised social goals developed with participants, self understanding plus emotional recognition and | Social skills necessary for developing and maintaining friendships and managing peer conflict and peer |

| | | | |
|---|---|---|---|
| | | expression goals | rejection |
| | Guiding principle | Cognitive behavior therapy (CBT), with special focus on functional analysis | CBT, especially focus on psychoeducation, role-play demonstration, and behavioural rehearsal |

Supplementary Table 2. Content for the Chinese 16-session version of KONTAKT$^{TM}$.

| Session | Participants | Group activities | Discussion topics | Homework |
|---|---|---|---|---|
| 1 | Children and parents | I am good at... | Presentation of KONTAKT$^{TM}$, review of confidentiality.<br><br>***Advanced Option*.**<br><br>How to introduce yourself.<br><br>Parent meeting: Presentation of KONTAKT$^{TM}$, Understanding autism, developing child's strengths, and talking about autism diagnosis with your child | Formulation of individual goals. |
| 2 | Children | Spin the bottle<br><br>***Optional.***<br><br>Story Cube[a] | Group rules<br><br>***Advanced Option*.**<br><br>How do I 'describe' myself in the group? How do I 'describe' the others in the group? | My supporters during KONTAKT$^{TM}$<br><br>Practising one of the child's midway goals |
| 3 | Children | What has changed?<br><br>***Optional.*** | What kind of child am I?[a]<br><br>***Advanced Option*.**<br><br>Sharing my social | A difficult situation at school, for example, when the child became angry at a classmate. |

| Session | Participants | Group activities | Discussion topics | Homework |
|---|---|---|---|---|
| | | The Wink game | difficulties with others.[a] | |
| 4 | Children | EU-Emotion<br>***Optional.***<br>Role-play, for example, conflict.[b] | How do gestures and facial expressions work?<br>***Advanced Option*.**<br>When do you use non-verbal communication? | Misunderstandings.<br>Children |
| 5 | Children | Role-play focusing on complex emotions.[b]<br>***Optional.***<br>What has changed? | Resolving misunderstandings.<br>***Advanced Option*.**<br>Understand jokes, irony, and sarcasm, and tell white lies. | Starting a conversation |
| 6 | Children | Treasure Hunt<br>**Optional.**<br>Role-play[b] | How to manage a new social situation?<br>***Advanced Option*.**<br>How can you support someone else in a new social situation? | Feeling lonely or not part of a group. |

| Session | Participants | Group activities | Discussion topics | Homework |
|---|---|---|---|---|
| 7 | Children | Talking to a stranger<br><br>**Optional.**<br><br>EU Emotion | Feeling lonely and being teased and/or bullied.<br><br>***Advanced Option***.<br><br>Managing difficult situations on social media. | Handling meeting cancellations or rejections.<br><br>Reviewing main and midway goals. |
| 8 | Children and parents | Charades[b]<br><br>**Optional.**<br><br>The Wink game | Choose a task to do during an excursion.<br><br>***Advanced Option***.<br><br>Talking to someone you like or want to be friends with.<br><br>Parent meeting:<br><br>Update on what has been covered on KONTAKT$^{TM}$ so far. | Prepare for the group excursion.<br><br>Individualised home assignments based on chosen tasks to be undertaken while on the group excursion. |
| 9 | Children | Group excursion, e.g., to a cafe, bubble tea shop, or convenience store.[a] | What to consider when going on an excursion? | Carrying out my task during the excursion. |
| 10 | Children | Video clip about non-verbal communication.[b] | Making/Canceling a time to meet up with someone | Talking to someone new or I do not know. |

| Session | Participants | Group activities | Discussion topics | Homework |
|---|---|---|---|---|
| | | **Optional.** Spin the bottle | ***Advanced Option***. How to stay in touch with a friend | |
| 11 | Children | Joint group activity **Optional.** Advanced treasure hunt[b] | Connecting with a person you do not know well ***Advanced Option***. Connecting through social media | Individualised homework assignments based on personal goals. |
| 12-15 | Children | Chosen by the selected Children. | Chosen by the selected Children. | Individualised homework assignments based on personal goals. |
| 16 | Children and parents | Jointly determined by the group | Reflection on what it was like to lead a KONTAKT$^{TM}$ session. Summarising the programme Parent meeting: Update on what has been covered on KONTAKT$^{TM}$ so far. | - |

[a]: Topics or scenarios different from the Australian English version of KONTAKT$^{TM}$ [2].

[b]: Activities that have been localised to match the Chinese culture.
