## Supplementary material for "Feasibility and cross-cultural validation of an adapted social skills group training programme (KONTAKT™ CHILD) for Chinese autistic children: a wait-list RCT protocol": online supplemental file 2

### Online Supplementary File 2: Assessment timelines and description for the measures

#### Tables

**Supplementary Table 3**. Schedule of enrolment, interventions, and data collection for the study

| | Involvement[a] | | | | | STUDY PERIOD | | | | | | |
|---|---|---|---|---|---|---|---|---|---|---|---|---|
| | | | | | | Enrolment | Allocation | Post-allocation | | | | |
| | C | P | T | E | F | | | | | | | |
| **TIME POINTS[b]** | | | | | | ***-T1*** | **0** | ***T1*** | ***T2*** | ***T3*** | ***T4*** | ***T5*** |
| **ENROLMENT:** | | | | | | | | | | | | |
| **Eligibility Screen** | | | | X | | X | | | | | | |
| **Informed consent** | | | | X | | X | | | | | | |
| **Allocation** | | | | | | | X | | | | | |
| **INTERVENTIONS (KONTAKT™):** | | | | | | | | | | | | |
| ***Immediate Intervention*** | X | X | X | X | X | | | ← | → | | | X |
| ***Delay Intervention*** | X | X | X | X | X | | | X | | ← | → | |
| **DATA COLLECTION ASSESSMENTS:** | | | | | | | | | | | | |
| ***Screening*** CBCL | | X | | | | X | | | | | | |
| ***Screening*** WISC-IV | X | | | X | | X | | | | | | |
| ***Screening*** ADOS-G | X | | | X | | X | | | | | | |
| ***Screening*** STI | | X | | X | | X | | | | | | |

| | | | | | | | | | | | | | |
|---|---|---|---|---|---|---|---|---|---|---|---|---|---|
| | SFI | X | X | | X | | X | | | | | | |
| | SES | | X | | | | X | | | | | | |
| ***Fidelity Outcomes*** | Recruitment | X | X | | | | X | X | | | | | |
| | Drop-out | X | X | | | | | | X | X | X | X | X |
| | Retention | X | X | X | | X | | | X | X | X | X | X |
| | Attendance | X | X | | | X | | | ↔ | ↔ | ↔ | ↔ | |
| | HCR | X | | | | X | | | ↔ | ↔ | ↔ | ↔ | |
| | Focus Group | X | X | | | X | | | | | X | | X |
| | KFC | | | | | X | | | ↔ | ↔ | ↔ | ↔ | |
| | GSRS | X | | | | | | | ↔ | ↔ | ↔ | ↔ | |
| | TFAQ | | X | | | | | | X | X | X | X | X |
| | NEQ | | X | | | | | | | | X | | X |
| ***Cost-effectiveness Outcomes*** | TIC-PC | | X | | | | | | X | X | X | X | X |
| | CHU9D | X | | | | | | | X | X | X | X | X |
| | EQ-5D-Y | | X | | | | | | X | X | X | X | X |
| ***Primary Efficacy Outcomes*** | CASS (+CRS) | X | | | X | | | | X | | X | | X |
| ***Secondary Efficacy Outcomes*** | SRS-2 | | X | X | | | | | X | | X | | X |
| | CRSES | X | | | | | | | X | | X | | X |
| | Koppitz DAPT | X | | | X | | | | X | | X | | X |
| | SCS-DAP | X | | | X | | | | X | | X | | X |

| | | | | | | | | | | | | | |
|---|---|---|---|---|---|---|---|---|---|---|---|---|---|
| | CORS | X | | | | | | | ← | → | ← | → | |
| | PRFQ | | X | | | | | | X | | X | | X |
| | ABAS-2 | | X | X | | | | | X | | X | | X |
| | PSS | | X | | | | | | X | | X | | X |
| | STARSS | X | | | X | | | | ← | → | ← | → | |
| ***Process Factors*** | TPOCS | X | | | X | | | | X | X | X | X | X |
| | PPKS | X | X | | | | X | | | | | | |
| | TASC | X | X | | | X | | | X | X | X | X | X |

a: Involvement: C: Child; P: Parent; T: Teacher; E: Evaluator blinded to the hypothesis; F: Trainer.

b: Time points: T1: Time 1; T2: Time 2; T3: Time 3; T4: Time 4; T5: Time 5.

Measures: ABAS-2:Adaptive Behaviour Assessment System-II; ADOS-G:Autism Diagnostic Observation Scale-Generic; CASS+CRS: Contextual Assessment of Social Skills + Conversation Rating Scale; CBCL: Child Behaviour Checklist; CHU9D: Child Health Utility 9D; CORS: Child Outcome Rating Scale; GSRS: Group Session Rating Scale; HCR: Homework Compliance Rate; KFC: KONTAKT™ Fedility Checklist; Kopptiz DAPT: Koppitz Draw-A-Person Test; NEQ: Negative Effects Questionnaire; PPKS: Preparedness for Participating KONTAKT™ Scale; PRFQ: Parental Reflective Functioning Questionnaire; PSS: Perceived school support; TASC: Therapeutic Alliance Scales for Children; TFAQ: Theoretical Framework of Acceptability Questionnaire; TIC-PC: Treatment Inventory of Costs in Patients with Psychiatric Disorders - Child Version; SCS-DAP: Self-Concept Scale-Draw-A-Person; SES: Socioeconomic Status Scale; SFI: Screening Face Interview; STARSS: Snack Time Assessment of Relationships and Social Skills; STI: Screening Telephone Interview; SRS-2: Social Responsiveness Scale-2; WISC-IV: Wechsler Intelligence Scale for Children-IV Chinese version.

### Description of measures

#### *Primary outcomes of the current protocol: Feasibility outcomes*

*Child Group Session Rating Scale (CGSRS)* is a 4-item visual analogue self-report scale measuring group-therapy alliance [1, 2]. Similar to the CORS, the child will rate each item by marking somewhere along the 10 cm visual analogue line. The GSRS will ask the child to respond about the relationship with both the group and therapists, the goals and topics of the session, the acceptability of the approach employed in the group, and the overall fit of the group. GSRS score ranges from 0 - 40, with a higher score indicating a better alliance. The GSRS has demonstrated adequate reliability (Cronbach's alpha > 0.80; test-retest reliability ranged from 0.42 to 0.62), construct and concurrent validity, and sensitivity to early change [2, 3]. Participants will rate the GSRS at the end of each session. The Cronbach's alpha will be used to assess the internal consistency of the CGSRS in the current trial.

*Theoretical Framework of Acceptability Questionnaire (TFAQ)* will be completed by parents at baseline, mid-point, and end-point. The TFAQ is a 10-item questionnaire measuring 10 different but correlated concepts of acceptability in healthcare interventions' design, evaluation, and implementation phases [4]. The concepts include affective attitude, burden, ethicality, intervention coherence, opportunity costs, perceived effectiveness, and self-efficacy. Following the instructions recommended by the authors of TFAQ, minor changes in wording were made according to the assessing period [4]. For example, the item about opportunity costs is stated as "Engaging in KONTAKT™ will interfere with my other priorities" at baseline assessment, while the same item is stated as "Engaging in KONTAKT™ has interfered with my other priorities." during the mid-point and end-point assessment period. Moreover, since parents will complete the TFAQ instead of the participants, parents will be asked to perceive the self-efficacy and opportunity costs of their children and their own. Parents will complete the TFAQ right before the first session of the KONTAKT™ commences, during the mid-point of the KONTAKT™ training, and right after the end of the KONTAKT™ training. In other words, both the Immediate Training Group (ITG) and the Delayed Training

Group (DTG) will complete this measure three times. A recent study employing the TFAQ to assess the social acceptability of the Senegalese government's measures against COVID-19 found the TFAQ to have good internal reliability (Cronbach's alpha > 0.80). [5]. The current study will also apply Cronbach's alpha to assess the internal consistency of the TFAQ.

*Negative Effects Questionnaire (NEQ)* is a 32-item measure designed to address the potential harms of psychological interventions [6]. The NEQ will be completed by parents. They will complete two versions of the NEQ, one related to their feelings and the other to their children (proxy). Each item is divided into three parts: (1) whether the negative effect in question exists; (2) if it exists, how severe the negative effect has been caused using a 5-point Likert scale; and (3) whether this effect is due to the psychological intervention or not. The NEQ has been previously applied in evaluating the Australian English version of KONTAKT™ [7, 8] and has been used in a study of SSGT intervention for Chinese adolescents [9], showing adequate psychometric properties (Cronbach's alpha = 0.83).

*Focus group semi-structured interviews* after completion of the KONTAKT™ programme and follow-up for the ITG will be conducted to explore the in-depth lived experiences of the participants, parents, and trainers. The qualitative data generated from these interviews can provide opportunities to capture factors influencing the relative efficacy of KONTAKT™ and support the further refinement of the KONTAKT™ manual. All focus groups will be conducted in person and hosted by experienced researchers not involved in the KONTAKT™ intervention. Each focus group will follow a similar topic guide, with a time length ranging from 30 to 60 minutes. Every discussion will be digitally recorded and transcribed verbatim in preparation for analysis.

#### *Secondary outcomes: preliminary efficacy*

##### Primary efficacy outcome

*Contextual Assessment of Social Skills (CASS)*, an objective and ecologically valid measure, is a videotaped role-play assessment of conversational skills. At the beginning of each role-play, the child will be introduced to an unfamiliar, similar-age, opposite-gender confederate. Next, they will be asked to act as if they are waiting for a recently joined new social group or club to start, and they can get to know each other. Then, the child will undergo two 3-minute semi-structured role-plays with different confederates, i.e., the interested and bored social context. Role-play contexts are manipulated by adjusting the confederate's interest level towards the conversation [10]. During the interested social context, the confederate will demonstrate interest and engage in the conversation. In contrast, the confederate will show his/her "boredom" using verbal and non-verbal gestures towards the conversation during the bored context. The child will first have the interested context and then the bored context. For each assessment time point, the child will be introduced to different confederates to ensure that the child will converse with an unfamiliar peer.

The confederates are undergraduates recruited through university advertisements and word-of-mouth. They will have a two-hour formal training on acting and practising their behaviours preceding the trial. In addition, they will be provided corrective feedback on recorded videos throughout the trial to ensure fidelity. Confederates will be unaware of the participant's group allocation, study hypotheses, and the scoring methods of the CASS.

To ensure the fidelity of administration, test administrators will follow the CASS operation manual developed by this article's first author (LU). For each role-play, the test administrator will read the identical instruction to the participant. After reading the instruction, the examiner will exit the room, and the role-play will start. After 3 minutes, the examiner will re-enter the room and announce the role-play to be ended. Subsequently, the child will complete the Conversation Rating Scale (CRS), a brief 5-item questionnaire regarding the confederate's interest in the conversation (perceived interest, friendliness, conversational flow, perceived distance, and sense of boredom) [10]. The child will rate on a 7-point Likert scale, with a higher total score indicating a higher perceived conversational interest in the corresponding social context. The CRS had a high internal consistency (alpha > 0.75) [10, 11].

The CASS includes nine coding items: Number of questions, number of topic changes, vocal expressiveness, gestures, positive affect, kinesic arousal, social anxiety, overall involvement, and overall quality of rapport. Except for the first two behaviour-counted items, the remaining seven items will be judged by performance quality or significant levels, rated on a scale of 1-7 (1 = low quality/very significant, 7 = high quality/no indication). A sum score is generated by adding all nine items, with a higher score indicating better social skills. Previous studies have demonstrated that the CASS is sensitive to intervention effects and has excellent internal consistency (alpha > 0.80) and acceptable inter-rater reliability (ICC > 0.50) [10, 11, 12].

The videos will be coded by trained coders achieving at least 70% consensus on six training recordings before rating the study videos. These coders will be trained by the authors of this article (ZH, BW, LU), who have already received training from the CASS developer Allison B. Ratto. Coders will be blinded to participants' allocation.

#### Secondary efficacy outcomes

##### Autistic presentation

*Social Responsiveness Scale-2 (SRS-2)* will be completed by parents and school teachers. The SRS-2 is a 65-item questionnaire assessing autistic traits across five correlated domains-social awareness, social communication, social cognition, social motivation, and autistic mannerisms [13, 14]. Parents and teachers will rate on a 4-point Likert scale (0–3), with higher total values indicating more significant autistic traits. The SRS-2 has been frequently applied to measure behaviour changes in autistic intervention [7, 15, 16] and demonstrates sensitivity to change and good psychometric properties in Chinese populations (Cronbach's alpha > 0.83) [9, 17]. In the current study, the raw scores of the SRS-2 will be used as recommended for intervention research.

#### Generalised Social skills

*Snack Time Assessment of Relationships and Social Skills (STARSS)* is adapted from the CASS and is used to evaluate the social interactions of KONTAKT$^{TM}$ participants during snack time, an important part of the KONTAKT$^{TM}$ sessions [18].

CASS focuses primarily on conversation-related social skills and behaviours in structured scenarios but lacks evaluation in areas beyond conversation skills, such as group dynamics and relationship building [10]. Also, CASS is based on a relatively structured environment, limiting the further inference of generalisation into broader social contexts. A recent adaptation of CASS called "Game Day" demonstrated the feasibility and potential value of using observational assessments in unstructured environments [19]. However, "Game Day" was time-consuming (lasting 40 minutes), and accurately counting specific indicators ("initiations" and "responses to initiations") was challenging during extended group video coding. Moreover, the authors suggested that shorter videos seemed to yield similar coding results. Therefore, STARSS was developed considering the structure of KONTAKT$^{TM}$ sessions, and the need for a simpler, naturalistic, and unstructured observational assessment.

#### Adaptive functioning

*Adaptive Behaviour Assessment System-II (ABAS-II)* will be completed by both parents and teachers. The ABAS-II is a frequently used and standardised tool measuring a child's everyday adaptive functioning across nine skill areas composing three domains [20]. Four domain composite scores are derived: General Adaptive Composite, Social, Practical, and Conceptual. The ABAS-II has been applied in studies of autism, including previous evaluation studies on KONTAKT$^{TM}$ [15, 16]. The Chinese version of ABAS-II has been standardised and demonstrated adequate psychometric properties (Cronbach alpha all $> 0.86$ in all subscales) [21].

#### Cognitive development and psychosocial well-being

*Child Outcome Rating Scale (CORS)* is an ultra-brief and child-friendly self-report measure assessing school-aged children's psychosocial functioning [22]. The child will answer four questions about himself/herself, his/her family, school, and overall perceived well-being. Below each item description is a 10 cm visual analogue line with a happy face on the left and an unhappy face on the right. The child will mark somewhere along each line to indicate his/her subjective feelings. The score of each item is generated by measuring the distance between the left end of the line and the mark that the child places on the line, ranging from 0 to 10. A total score of the CORS is generated by summing up all four items. Previous psychometric studies have indicated that the CORS has adequate reliability (Cronbach's alpha = 0.84) and construct validity [22]. Moreover, the CORS was sensitive to changes due to intervention, thus suitable for measuring children's self-report psychosocial functioning treatment outcomes [23]. Participants will rate the CORS before each session starts. As the Chinese normative sample data is lacking, the current study will use Cronbach's alpha to assess the reliability of this measure.

*Koppitz Draw-A-Person Test (KDAPT)* is a psychological projective device in which the child will be instructed to draw three depictions of people - a man, a woman, and himself/herself [24, 25]. Although the KDAPT can be administered in groups, the current study will implement the test individually.

According to Koppitz, children's drawings reflect transient attitudes and emotional indicators [24]. Therefore, human figure drawings (HFD) can measure mental and socio-emotional maturity. One previous study has shown that KDAPT is effective in assessing negative emotions in Chinese school-aged children, demonstrating good discrimination validity in emotional distress and concurrent validity [26], however normative data has yet to be established.

The child will be instructed to seat at the desk and make three drawings, one for each A4 sheet, on three sheets of paper using a 2B pencil. Each time before the child draws a depiction, the test administrator will give the following instructions to the child to ensure he/she will draw a full-body portrait:

"Please draw as carefully as you can. It can be any person you want to draw, but remember to draw the whole man/woman/yourself from the top of the head to the toe, and do not draw the caricature or the match man."

The child can then start to draw. Rubber will also be provided in case the child wants to correct the drawings. Only one sheet of A4 paper is provided for each portrait drawn.

The KAPT consists of two sets of indicators, i.e. non-verbal cognitive development and emotional indicators [24]. The scoring system proposed by Kopptiz can estimate the mental development level of 5 to 12-year-olds from human figure drawing (HFD). The scoring system will calculate the expected and unexpected developmental indicators with the corresponding age. Each expected indicator is assigned a value of 0 (not missing) and -1 (missing). On the other hand, each unexpected indicator will be assigned a value of 0 (not present) and +1 (present). +5 is routinely assigned as the baseline score in the final sum score calculation to avoid negative scores. The sum score can be converted into a range estimate of the level of mental development [27].

Another set of indicator system developed by Koppitz is the 38-item emotional indicators. The 38 emotional indicators, which are uncommon in "healthy" children's HFD (<16%), are not related to a child's age or maturity but reflect anxiety, worries and attitudes. Examples of these indicators include shading, absence of particular parts of the body, abnormal figure size, detailing, and asymmetry. Each emotional indicator will be coded as either 0 (absence) or 1 (present). A total score will be generated by summing all items.

Two trained coders will code these drawings independently. In order to facilitate the coding process and enhance reliability, the administration and the coding will follow an operation manual developed explicitly for this study. The trained coders and test administrators will be blinded for participants' allocation statuses and assessment period, and the Kappa coefficient will be performed to assess the inter-rater reliability. The KDAPT has acceptable test-retest reliabilities from 0.81-0.91 in previous studies [25, 28, 29].

#### Self-esteem and self-concept

*Self-concept Scale-Draw-A-Person (SCS-DAP)* is a quantified self-concept scale measuring children's self-concept via projective technique [30]. The child will be instructed to draw his/her whole body using paper and pen (see above the KAPT for the implementation procedure). The SCS-DAP contains nine coding items: Reinforcement, Erasures, Sketchy Lines, Immaturity, Incompleteness, Opposite Sex Identification, Transparency, Primitiveness, and Distortion. Each item will be rated on a Likert 5-point scale, from 5 (markedly absent/0-20%) to 1 (markedly present/81-100%). The total score of the SCS-DAP ranges from 9 to 45, with a higher score indicating a better self-concept. Two blinded coders will code the SCS-DAP. Previous studies suggested that the SCS-DAP has good face and concurrent validity (Pearson correlation with clinical interview judgement of self-concept was 0.64), excellent inter-rater reliability (Kendall's Coefficient of Concordance W = 0.819) [31] and acceptable internal reliability [30, 32] in developmental disabilities children. The current study will use the Kappa coefficient to assess inter-rater reliability, and Concrach's alpha to assess internal reliability.

*Child Rosenberg Self-esteem Scale (CRSES)* is a recently developed self-reported 10-item scale assessing global self-esteem (SE) in children aged 7-12 [33]. The CRSES is a modified version of the Rosenberg Self-esteem Scale (RSES), the most widely applied and reliable measure for describing SE. Compared with the RSES, The CRSES utilised a simpler phraseology that school-aged children can understand. For example, the first item of RSES ("I feel like I am a person of worth, at least on an equal plane with others") was simplified into "I feel that I'm as good as everyone else". Children will rate each item using a 4-point Likert scale (1= very true, 4= definitely not true), with a total score ranging from 10-40. The scale consists of half positive and negative statements, with the negative items reversely scored. Therefore, a higher total score implies a better perceived SE. Results of the psychometric study supported the CRSES has convergent and construct validity, and the internal reliability is acceptable (Cronbach's alpha = 0.71) [33]. In the present study, the CRSES was translated into Mandarin Chinese and checked for readability using the Chinese Readability Index Explorer 3.0 (CRIE3.0) [34]. The current study will apply Cronbach's alpha to assess the internal reliability of the CRSES.

#### Parental reflective functioning

*Parental Reflective Functioning Questionnaire (PRFQ)* is a relatively brief self-report assessment for parental reflective functioning [35]. Parental reflective functioning (PRF), or mentalising, refers to the parent's capacity to reflect on his/her inner mental state experiences and those of his/her child [36]. PRF is a predominant feature of adaptive parenting, especially in fostering efficacy in dealing with distressing interactions [37, 38]. PRF is positively related to the child's cognitive and social development [39, 40]. Therefore, parents with better PRF may better facilitate their children to engage in the interventions. The revised Chinese version of PRFQ includes 12 items (PRFQ-12C), assessing three dimensions of PRF, viz. Pre-mentalisation (PM), Certainty about mental states (CMS), and Interest in and curiosity about mental states (IC) [41]. The PM dimension reflects that the parent struggles to understand the subjective world of his/her child, with a tendency to make maladaptive and malevolent attributions. The CMS dimension measures how much confidence the parent has in understanding his/her child's mental states, with an overly high score reflecting a lack of recognition of the opacity of mental states (hypermentalising) and an overly low score reflecting an almost complete lack of certainty on the mental states (hypomentalising). Finally, the IC dimension addresses the degree of the parent's genuine curiosity about the mental states underlying his/her child's observable behaviours. Similar to the CMS dimension, a low score on the IC dimension indicates an almost absence of interest in the child's mental states, and an overly high score reflects intrusive hypermentalising. The parent will rate the PRFQ using a 7-point Likert scale, from 1 (strongly disagree) to 7 (strongly agree). The PRFQ contains two reverse scoring items. A total mean score and three domain subscale mean scores will be generated. The Chinese version of PRFQ has shown to have acceptable to good internal consistency (Omega coefficients ranged from 0.68-0.82), good discriminant validity (HTMT ranged from 0.18-0.59), good construct validity, and acceptable convergent validity [41].

##### Perceived school support

Parents will rate the child's perceived school support level on a 10-point Likert scale, from 1 (very low) to 10 (very high).

#### Additional outcome

##### CASS during Snack Time (STARSS)

In order to examine the generalization of social skills into a naturalistic setting with peers, children's performance during the Snack Time in KONTAKT sessions (early and late stage of the KONTAKT) will be analysed using a modified version of CASS (Snack Time assessment of relationships and social skills, STARSS). The development of STARSS referred to the Game Day, another modified version of CASS, which is also an assessment of social skills in a group-setting.

During snack time, trainers encourage participants to engage in prosocial behaviours, such as helping others or asking if anyone wants anything, before formally starting snack time. Children have approximately 5 minutes for snack time, during which trainers step aside and observe the interactions. Trainers should minimise participating in conversations and interactions, only stepping in when necessary, such as in the case of clear verbal or physical conflicts between participants or the need to clean up spilt food or drinks promptly to avoid disrupting the rest of the class.

In STARSS, additional scoring items include "responding to peers' topics or comments" and "negotiation" in addition to CASS items. Furthermore, as STARSS emphasizes relationships, the Overall Quality of Rapport will pay special attention to whether children engage in reciprocal behaviours to foster relationship building (e.g., helping peers prepare and obtain snacks, inviting peers to share snacks).

Three video recordings are selected for scoring, representing the beginning, middle, and end of the programme. Specific sessions are chosen randomly, excluding the first session, the ninth session (Excursion), and the sixteenth session. Evaluators are unaware of the programme stage during which the video was recorded, and independent dual scoring is performed.

### *Process Factors*

#### Alliance

*Therapeutic Alliance Scale for Children (TASC)* will be completed by both therapists, children and parents. The TASC is a 12-item inventory assessing alliance with three different respondent versions [42, 43]. Respondents will rate each item using a 4-point scale to reflect their agreement towards the statement, with higher points for higher agreement. The TASC total score ranges from 12-48; a higher score means a better alliance. The TASC has been previously applied in SSGT interventions for autistic children and showed excellent internal consistency (Cronbach's alpha = 0.87) [44]. The study also showed that the baseline and the change score of TASC were predictive of treatment outcome. In the current study, Cronbach's alpha will be applied to assess internal reliability.

#### Readiness for change

*Preparedness for Participating in KONTAKT™ Scale (PPKS)* is a 3-item visual analogue scale that assesses children's and parents' readiness to participate in KONTAKT™. The PPKS is an adopted version initially developed by Albaum et. al., 2020 [45]. Each child and his/her parent will be asked at the intake interview the following three questions: (1) How much do you want to be part of the programme? (2) How much do you want yourself/your child to change? (3) How hard are you willing to work when you (and your child) participate in KONTAKT™? The respondent will answer the questions on an 8-point visual analogue scale, ranging from 0 (not at all) to 8 (very, very much). A total score summing up all three item ratings, ranging from 0 to 24, will be used instead of averaging the three ratings. The original scale developed by Albaum et. al., 2020 [45] did not provide psychometric information of validity and reliability. In the current study, we will apply Cronbrach's alpha to assess internal consistency.

#### *Cost-effectiveness outcomes*

##### Economic impact

*Child Health Utility 9D (CHU-9D)* is a widely used generic preference-based health-related quality of life (PBHRQOL) self-reported measure for children and adolescents [46, 47]. It comprises nine dimensions: sad, worry, pain, tiredness, annoyance, school, sleep, daily routine, and activities. The child rates his/her feelings "today" on a 5-level response, from no problems (1) to experiencing many problems (5). Each level of an item has a corresponding severity brief statement to aid understanding. The CHU-9D estimates the child's quality-adjusted life years (QALYs). The raw total score of the CHU-9D will be transferred to a universal score using the validated Chinese scoring algorithm, with a score ranging from 0 (worst) to 1 (best) [47]. The Chinese version of CHU-9D had acceptable internal consistency (Cronbach's alpha = 0.77), moderate test-retest reliability (ICC = 0.65), and good construct and convergent validity [47].

*EQ-5D-Y* is one of the most frequently used generic HRQOL measures for children and adolescents [48, 49]. The EQ-5D-Y derives from the EQ-5D, containing five health dimensions: mobility, self-care, usual activities, having pain or discomfort, and feeling worried, sad, or unhappy. Each dimension has three functioning levels (no, some, and a lot of problems). Besides, the EQ-5D-Y include one visual analogue scale (EQ VAS) ranging from 0 -100, where the end-points are labelled as "the best health you can imagine" and "The worst health you can imagine". The current study will apply the proxy version and the validated Chinese scoring algorithm of the EQ-5D-Y [50]. The Chinese proxy version of EQ-5D-Y has demonstrated good to excellent reliability (ICC for EQ VAS = 0.833, test-retest reliability for EQ-5D-Y dimensions using Gwet's agreement coefficient ranged from 0.332-0.668) [51].

##### Healthcare consumption and productivity loss

*The tailored version of the treatment inventory of costs in patients with psychiatric disorders - child version (TIC-PC)* will be applied in the present study to measure healthcare-related consumption and productivity loss [52, 53]. The tailored version of TIC-PC has been used in the previous evaluation study of the Australian English version of KONTAKT™ [54]. The questionnaire consists of six sections: healthcare visits, medications and supplements, seeking medical care or treatments, school support, and productivity losses suffered by the target child and parents. The TIC-PC psychometric properties have not been published yet, while TIC-P has shown adequate test-retest reliability, feasibility, and construct validity [52]. As assessing these properties is not feasible in this study, we will focus on feasibility by evaluating the TiC-P response rate and data completeness of the questionnaire items. Additionally, we will assess the completeness of reported medication usage patterns, including medication names, doses per intake, number of doses per day, and total days medications were taken in the past 60 days.
