## Supplementary material for "Feasibility and cross-cultural validation of an adapted social skills group training programme (KONTAKT™ CHILD) for Chinese autistic children: a wait-list RCT protocol": online supplemental file 3

### Online Supplemental File 3: Supplementary information on the cost-effectiveness analysis of the Chinese KONTAKT$^{TM}$ CHILD study

The healthcare perspective will be considered from the particular interest of service providers, participants, and parents. The societal perspective will acquire the healthcare service costs, transportation fees to the healthcare services, additional education costs due to the child's diagnosis of autism, parents' out-of-pocket costs (e.g., home accommodation due to the child's diagnosis of autism), parental and the child's productivity losses due to autism, and unpaid caregiver time (additional hours of care provided to the child) using the TIC-PC [1]. Productivity losses of the parents are divided into absenteeism and presenteeism [2, 3]. Absenteeism is quantified by multiplying the number of days off work due to the child's health condition by the hours per workday of the parent. This will then be multiplied by the parent's salary. The calculation of presenteeism productivity loss will apply this formula: number of impaired workday x [1 - (efficacy score/10)] x number of hours per workday, in which the efficacy score ranges from 0 (present at work but no function at all) to 10 (bothered, but can function equally as a typical day) [1, 2, 3]. Unpaid caregiver time will apply to the price paid for equal care time provided by a home care worker. For prescription and non-prescription drugs, the unit costs are obtained from the Guangdong Provincial Bureau of Medicine website [4].
